# Unbiased sequencing of *Mycobacterium tuberculosis* urinary cell-free DNA reveals extremely short fragment lengths

**DOI:** 10.1101/2021.06.22.21258723

**Authors:** Amy Oreskovic, Adam Waalkes, Elizabeth A. Holmes, Christopher A. Rosenthal, Douglas P.K. Wilson, Adrienne E. Shapiro, Paul K. Drain, Barry R. Lutz, Stephen J. Salipante

## Abstract

Urine cell-free DNA (cfDNA) presents an attractive target for diagnosing pulmonary *Mycobacterium tuberculosis* (TB) infection but has not been thoroughly characterized. Here, we aimed to investigate the size and composition of TB-derived urine cfDNA with minimal bias using next-generation DNA sequencing (NGS). To enable analysis of highly fragmented urine cfDNA, we used a combination of DNA extraction (Q sepharose) and single-stranded sequence library preparation methods demonstrated to recover short, highly degraded cfDNA fragments. We examined urine cfDNA from ten HIV-positive patients with confirmed pulmonary TB (nine of which had TB cfDNA detectable by qPCR) and two TB-negative controls. TB-derived cfDNA was identifiable by NGS from all TB-positive patients. TB urine cfDNA was significantly shorter than human urine cfDNA, with median fragment lengths of ≤19–52 bp and 42–92 bp, respectively. TB cfDNA abundance increased exponentially with decreased fragment length, with a peak fragment length of ≤19 bp in most samples. Our methodology also revealed a larger fraction of short human genomic cfDNA than previously reported, with peak fragment lengths of 29–53 bp. Urine cfDNA fragments spanned the TB genome with relative uniformity, but nucleic acids derived from multicopy elements were proportionately overrepresented, providing regions of inherent signal amplification beneficial for molecular diagnosis. This study demonstrates the potential of urine cfDNA as a diagnostic biomarker for TB and will inform improved design of TB urine cfDNA assays. Methods capable of targeting the shortest cfDNA fragments possible will be critical to maximize TB urine cfDNA detection sensitivity.

## INTRODUCTION

There is a critical need for diagnostics for pulmonary *Mycobacterium tuberculosis* (TB) infection that do not rely on the collection of sputum samples, which are difficult to obtain from many patients. Even when sputum is available for testing, existing sputum-based TB assays have reduced sensitivity for diagnosis of paucibacillary, HIV-associated, pediatric, and extrapulmonary TB (1–3). Transrenal urine cell-free DNA (cfDNA) is a promising, easy-to-collect biomarker for TB with the potential to diagnose TB across these key underserved patient populations (4–9), but it has not yet been extensively characterized or validated as a diagnostic analyte.

In particular, the fragment length distribution of transrenal microbial cfDNA, including TB cfDNA, has not yet been robustly determined. Recent work employed NGS sequencing of three TB urine cfDNA samples using conventional methods and identified the presence of short fragments (19–44 bp) but provided minimal characterization of the overall fragment length distribution (10). Moreover, conventional methods for DNA extraction and next-generation sequencing (NGS) may systematically underestimate the proportion of short urine cfDNA molecules present because they have poor retention of degraded DNA fragments. In contrast, single-stranded NGS library preparation (11, 12) improves yield of short cfDNA fragments (<100 bp) and reveals the presence of highly degraded forms of cfDNA, including microbial cfDNA, that are less protected from nuclease digestion than human genomic cfDNA (13, 14).

In this study, we aimed to better characterize the fragment length distribution of TB-derived urine cfDNA using NGS and identify any potentially over-represented sequences suitable for targeting in diagnostic assays. In addition to using single-stranded library preparation to improve the recovery of highly degraded DNA fragments, we selected a DNA extraction method based on Q Sepharose anion exchange resin known to increase retention of short urine cfDNA relative to other urine cfDNA extraction methods, including commonly-used commercial extraction kits (15, 16). We theorized that this combination of methods would minimize biases resulting from fragment length and enable more accurate NGS characterization of TB urine cfDNA.

## MATERIALS & METHODS

### Participant enrollment and urine collection

Participants were enrolled at Edendale Hospital in Pietermaritzburg, South Africa between October 2019 and February 2021 (approved by the University of KwaZulu-Natal Biomedical Research Ethics Committee, #BE475/18). Adults (≥16 years old) with a positive admission sputum Xpert MTB/RIF Ultra (Cepheid, Sunnyvale, CA, USA) or adults with HIV, regardless of reason for admission, were recruited and provided sputum for Xpert Ultra testing. Patients with >24 hours of anti-TB treatment were excluded. All participants provided written informed consent.

Participants provided a urine sample (50–200 mL) at the time of enrollment and/or an early morning, first-void sample the morning after enrollment. The samples were not obtained mid-stream. Immediately after collection, urine was mixed in 10 mL aliquots with EDTA to a final concentration of 25 mM and Tris-HCl pH 7.5 to a final concentration of 10 mM. Urine was stored in DNA LoBind tubes (Eppendorf, Hamburg, Germany) at -80°C until shipping, shipped on dry ice to the University of Washington, and stored at -80°C until processing. Samples were de-identified prior to testing.

### Clinical data, sputum testing, and urine lipoarabinomannan (LAM) testing

Clinical data were collected as follows for all participants: gender, presence of TB symptoms (cough, fever, night sweats, weight loss, loss of appetite, fatigue), TB treatment duration, HIV test result, and CD4^+^ cell count (for participants living with HIV only). Expectorated sputum was submitted to the South African National Health Laboratory System (NHLS) for Xpert MTB/RIF Ultra testing (if not already performed) and confirmatory solid and liquid mycobacterial culture. Mycobacterial culture was performed at the NHLS Provincial TB Reference Laboratory using both Middlebrook 7H11 solid agar medium and the liquid BACTEC mycobacterial growth indicator tube (MGIT) 960 system (BD, Franklin Lakes, NJ, USA) for each sputum sample. Cultures were incubated for up to 42 days. Culture plates were read at 3 and 6 weeks, and *M. tuberculosis* was identified from solid or liquid cultures using niacin and nitrate testing. Participants were considered culture-positive with growth from either the solid or liquid culture. Urine (60 µL) was tested using Alere Determine TB LAM Ag (Abbott Laboratories, Chicago, USA). Participants were categorized as TB-positive if at least one of the Xpert MTB/RIF Ultra or mycobacterial culture were positive. Participants were categorized as TB-negative if neither Xpert MTB/RIF Ultra or culture were positive and no clinical diagnosis of TB was assigned within 2 months of enrollment.

### cfDNA extraction using Q Sepharose anion exchange resin

Urine was thawed at 37°C and centrifuged for 5 minutes at 8,000g to pellet cell debris. The cell-free urine supernatant was transferred to new 15 mL DNA LoBind tubes (Eppendorf). cfDNA was extracted using a previously-described Q Sepharose extraction protocol (15, 16), with some modifications. Specifically, we increased the spin speed of the silica column wash steps from 800g to 8,000g to avoid column clogging that occurred when processing some clinical samples, reduced the elution volume from 106 µL to 50 µL to increase the concentration of eluted cfDNA prior to library preparation, and reduced the volume of eluate per PCR well from 5 µL to 2 µL to avoid PCR inhibition. We found that these modifications had no effect on the total cfDNA yield or recovery of a spiked 50 bp positive control sequence (not shown). Each 10 mL urine sample was mixed with 300 µL Q Sepharose Fast Flow resin (GE Healthcare, Waukesha, WI) and rotated at room temperature for 30 min. The resin was pelleted for 5 minutes at 1,800g, resuspended in 1 mL low salt buffer (0.3 M LiCl, 10 mM NaOAc pH 5.5), transferred to a Mini Bio-Spin Column (Bio-Rad Laboratories, Hercules, CA), and filtered for 1 minute at 800g. The resin was washed four times with 0.5 mL low salt buffer for 30 seconds at 800g. DNA was eluted using 670 µL high salt buffer (2 M LiCl, 10 mM NaOAc pH 5.5) for 3 minutes at 800g. The eluate was mixed with 2 mL 95% ethanol and applied in 700 µL increments to a QIAquick column (Qiagen, Hilden, Germany) with 30 seconds centrifugation at 8,000g for each increment. The column was washed twice with 0.5 mL 2 M LiCl in 70% ethanol and twice with 0.5 mL 75 mM KOAc pH 5.5 in 80% ethanol for 30 seconds at 8,000g. The column was dried for 3 minutes at 20,000g and DNA was eluted in 50 µL elution buffer (Qiagen) for 2 minutes at 20,000g.

### Quantification of cfDNA

The total concentration of purified cfDNA was measured using the Qubit dsDNA HS Assay Kit (Thermo Fisher Scientific, Waltham, MA, USA). The concentration of *Mycobacterium tuberculosis* (MTB) complex-specific cfDNA was measured using qPCR of a 40 bp region of the variable copy number insertion sequence IS6110. PCR was carried out in triplicate for each sample, with 2 µL eluate amplified in a 50 µL reaction containing 1.25 U OneTaq Hot Start DNA Polymerase (New England Biolabs [NEB, Ipswitch, MA, USA]), 1X OneTaq GC Reaction Buffer (NEB; 80 mM Tris-SO_4_, 20 mM (NH_4_)_2_SO_4_, 2 mM MgSO_4_, 5% glycerol, 5% dimethyl sulfoxide, 0.06% IGEPAL CA-630, 0.05% Tween-20, pH 9.2), 0.8 mM dNTPs (NEB), 0.4X EvaGreen (Biotium, Fremont, CA, USA), 200 nM forward primer (5’-CGAACCCTGCCCAGGTCGA-3’), and 200 mM reverse primer (5’-GTA+GCAGA+CCTCACCTATGTGT-3’, where “+G” and “+C” indicate locked nucleic acid bases). Primers were ordered from Integrated DNA Technologies (Coralville, IA, USA) with HPLC purification. qPCR was carried out in a CFX96 Touch Real-Time PCR Detection System (Bio-Rad Laboratories, Hercules, CA, USA) using an initial incubation period of 94°C for 3 minutes, followed by 45 amplification cycles of 94°C for 30 seconds, 64°C for 30 seconds, and 68°C for 1 minute. Quantification cycle (C_q_) values were determined using the CFX Maestro software version 1.1 (Bio-Rad Laboratories) at a threshold of 500 RFU and recovered copies were calculated using a standard curve. PCR products were confirmed by post-amplification melt curve analysis from 65°C to 95°C in 0.5°C increments every 5 seconds.

### Next-generation sequencing

Sequencing libraries were prepared with 1 to 45 ng purified cfDNA using the SRSLY method as described elsewhere (12), or using the commercially available formulation of that protocol, SRSLY PicoPlus kit (Claret Biosciences), with some modifications. Specifically, to retain low molecular weight cfDNA fragments, Monarch Genomic DNA Purification Kit (NEB) was used for purification of library fragments after the phosphorylation/ligation reaction, final library purification was performed using 1.8X volumes AMPure XP beads (Beckman Coulter), and size selection of the final library was not performed. Two to 11 replicates were generated per specimen. Sequencing was carried out using a NextSeq500 (Illumina) with 150bp Paired-End chemistries.

### Data analysis

Sequence reads from replicate library preparations of a given specimen were combined prior to data analysis. Reads were trimmed using the fastq-mfc algorithm of ea-utils-1.1.2.779 (17), with minimum post-trimmed sequence length set to 15 bp. The taxonomic composition of processed reads was cataloged using kraken2 (18). Individual read pairs classified as human (downsampled to 200,000 reads) or as members of the *Mycobacterium* genus were isolated for further analyses. Sequence read mapping was performed against the human genome (hg38) or the *Mycobacterium tuberculosis* H37Rv genome (GenBank Accession AL123456.3), respectively, using bwa-mem (v0.7.12) (19) with default parameters. Sequence reads with a minimum mapping quality score of 5 or greater were retained for further analysis. This approach was ultimately able to identify fragments ≥19 bp in length, equivalent to the default minimal seed length required by bwa-mem. cfDNA fragment length distributions were determined using deepTools (20), with the “distanceBetweenBins” flag set to 100.

For studies of multicopy elements IS6110 and IS1081, reads were mapped directly to those reference sequences (GenBank Accession X17348.1 and X61270.1, respectively) using bwa-mem as described above, and read counts were subsequently quantified. All statistical analysis was conducted using GraphPad Prism v8.1.2 (San Diego, CA, USA) with a significance level of 0.05.

### Data availability

Reads mapping to the TB genome are available from the NCBI Sequence Read Archive (SRA) under accession PRJNA725220.

## RESULTS

### Q Sepharose DNA extraction and cfDNA quantification

We extracted cfDNA from the urine of 29 TB-positive and 5 TB-negative participants enrolled at Edendale Hospital in Pietermaritzburg, South Africa. We detected TB-specific cfDNA by Q Sepharose extraction and IS6110 qPCR in 14/29 (48.3%) of urine samples from TB-positive participants and 0/5 (0%) of urine samples from TB-negative participants. A summary of the detected concentrations of total cfDNA and TB-specific cfDNA is given in Table 1.

**Table 1:**
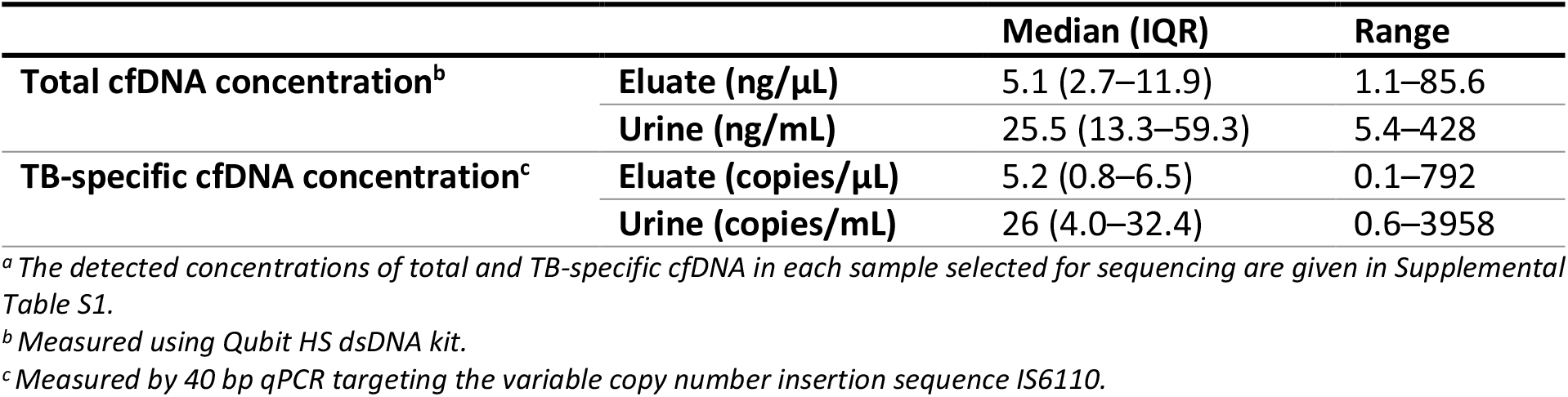
Concentrations of total and TB-specific urine cfDNA detected after Q Sepharose extraction.^a^.

|  |  | Median (IQR) | Range |
| --- | --- | --- | --- |
| Total cfDNA concentration<sup>b</sup> | Eluate (ng/μL) | 5.1 (2.7–11.9) | 1.1–85.6 |
|  | Urine (ng/mL) | 25.5 (13.3–59.3) | 5.4–428 |
| TB-specific cfDNA concentration<sup>c</sup> | Eluate (copies/μL) | 5.2 (0.8–6.5) | 0.1–792 |
|  | Urine (copies/mL) | 26 (4.0–32.4) | 0.6–3958 |

<sup>a</sup> The detected concentrations of total and TB-specific cfDNA in each sample selected for sequencing are given in Supplemental Table S1.
<sup>b</sup> Measured using Qubit HS dsDNA kit.
<sup>c</sup> Measured by 40 bp qPCR targeting the variable copy number insertion sequence IS6110.

### Urine cfDNA sequencing

We selected nine TB-positive samples with the highest concentrations of TB-specific cfDNA detectable by IS6110 qPCR, one TB-positive sample with no detectable TB-specific cfDNA by IS6110 qPCR, and two TB-negative samples for single-stranded library preparation and sequencing. Demographic and clinical data for the subset of participants whose urine samples were selected for sequencing are given in Supplemental Table S1. All participants were HIV-positive with a median CD4 count of 141 cells/mm^3^ (IQR: 59–516 cells/mm^3^). Participants were 42% female and 58% male. 50% of TB-positive participants had a positive urine LAM result.

A total of 30 million to 113 million sequence reads were generated per specimen, resulting in 24 to 99 million reads after initial quality filtering to computationally remove self-ligated adaptor sequences (Supplemental Table S2). Library complexity, the measured proportion of unique sequence fragments sequenced per library, was high for all cases, ranging from 95.2% to 100% per specimen.

### Urine cfDNA taxonomic composition

We initially characterized the taxonomic composition of cfDNA reads using metagenomic analysis techniques (18). A summary is provided in Supplemental Table S2 and the full output of the metagenomic analysis is given in Supplemental Dataset S1. For all cases, the majority of quality-filtered sequence reads (84.5–99.2%) corresponded to human nucleic acid, while the next most abundant taxa were attributable to microorganisms comprising the normal skin or genitourinary microbiota, primarily species within *Actinobacteria, Proteobacteria*, and *Bacteroidetes* (21). The proportion of reads originating from bacteria of any kind averaged 1.71% per case (range 0.57% to 3.69%), with no difference in bacterial sequence load in comparing between TB-positive and TB-negative study participants (p=0.71 by 2-tailed T-test). The remainder of reads were distributed among higher level taxonomic classifications, viral and phage sequences, and unclassified reads.

Sequences putatively classified as originating from TB or human were first mapped against their respective reference genomes to confirm their identity prior to further analysis. Following this quality control step, some number of TB-derived reads were identified in all specimens from TB-positive participants (10/10 specimens), including the one patient lacking qPCR-detectable TB IS6110 cfDNA (Supplemental Table S2). Significantly, no reads mapping to the TB genome were identified from either of the TB-negative individuals (0/2 specimens). An average of 2,332 reads originated from the TB genome in TB-positive patients (range 4 to 19,547 reads per case), corresponding to a proportion of 0.0000100% to 0.0201% of total reads.

### Human and TB urine cfDNA fragment lengths

We next used sequencing data to explore the length of cfDNA fragments derived from the human and TB genomes (Figure 1).

**Figure 1:**
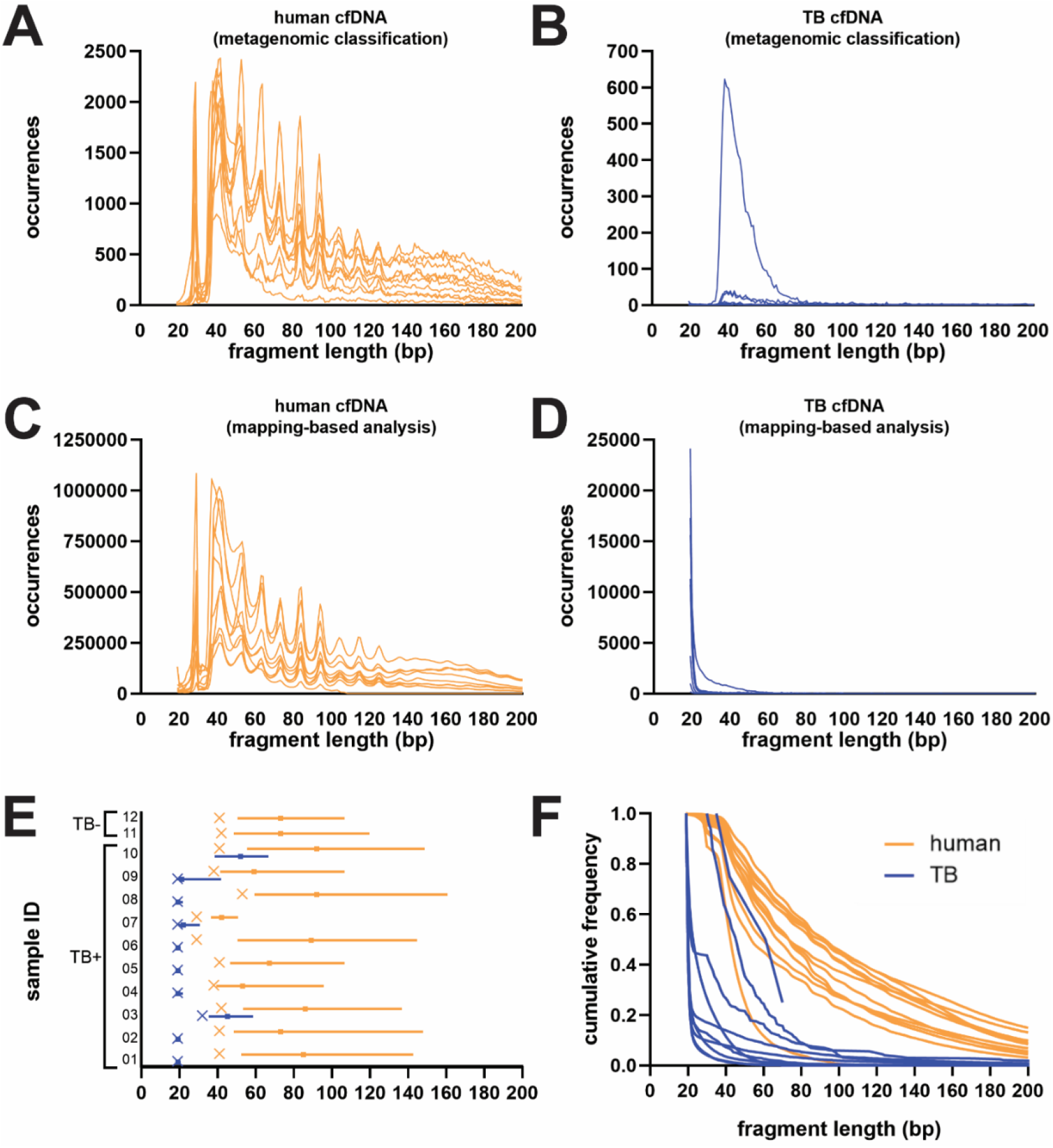
TB urine cfDNA is significantly shorter than human genomic urine cfDNA. **(A)** Fragment length distributions of urine cfDNA in each sample classified as human by metagenomic analysis techniques (n=12). **(B)** Fragment length distributions of urine cfDNA in each sample classified as TB by metagenomic analysis techniques (n=10). **(C)** Fragment length distributions of urine cfDNA in each sample mapped to the human genome (n=12). Individual plots for each sample are given in Supplemental Figure S1. **(D)** Fragment length distributions of urine cfDNA in each sample mapped to the TB genome (n=10). Individual plots for each sample are given in Supplemental Figure S2. **(E)** Characterization of fragment length for cfDNA mapped to the TB genome (blue, n=10) and human genome (orange, n=12) in each sample. Bars indicate median fragment length and IQR. “x” indicates mode fragment length. No mode length is shown for Sample 10 because it was multimodal with a low number of reads mapped to TB. The median, IQR, and mode fragment length for each individual sample are given in Supplemental Table S1. **(F)** Cumulative frequency of TB (blue, n=10) and human genomic (orange, n=12) cfDNA by fragment length in each sample.

Human cfDNA showed a relatively broad distribution of fragment lengths, with the abundance of reads inversely proportional to fragment length (Figure 1A). A periodicity in fragment abundance occurred at approximately 10 bp intervals, as expected based on nucleosome length and the results of previous cfDNA sequencing studies (14, 22–25). The most abundant fragment length for human urine cfDNA ranged from 28–53 bp across samples (Supplemental Table S1). The median fragment lengths for human urine cfDNA were 45–97 bp (Supplemental Table S1).

The abundance of TB urine cfDNA similarly increased with decreasing fragment length. However, in contrast to human cfDNA, TB-derived cfDNA displayed no periodicity and showed a left-shifted distribution (Figure 1B). In samples with enough reads to determine the peak TB cfDNA fragment length, it ranged from 38–43 bp (Supplemental Table S1). The median fragment lengths for TB-derived urine cfDNA were 39–97 bp (Supplemental Table S1) and were significantly shorter than those for human cfDNA (p=0.02 by Wilcoxon matched pairs test).

Although this analysis recovers TB-derived cfDNA with high specificity, a drawback is that it preferentially identifies longer sequences, which have a correspondingly higher probability of containing sequence motifs that uniquely identify them as TB. Shorter fragments that legitimately derive from the TB genome are more likely to share significant similarity with other species by homology or by chance alone, and will consequently be excluded.

To provide an analysis that is less biased with respect to sequence length, we next aligned all sequence reads to the TB and human reference genomes, regardless of their metagenomic classifications, and retained for analysis those that could be successfully mapped. A comparatively greater number (average 22,545; range 4 to 78,240) and proportion (average 0.027%; range 0.000010% to 0.081%) of reads matching the TB genome were recovered from TB-positive patients (Supplemental Table S2). A small number of reads from TB-negative participants also mapped to the TB genome, suggesting minor, artifactual contributions of cfDNA from other organisms that have been mapped to the TB genome. Nevertheless, the proportions of TB-mapped reads from negative patients (0.0000318% and 0.000178% of reads, corresponding to read counts of 9 and 42, respectively) were three orders of magnitude less than the average for TB-positive patients, despite the two groups having comparable proportions of total bacterial cfDNA by metagenomic analysis. Moreover, we found no correlation between the proportion of reads mapping to the TB genome and the proportion of total bacterial reads cataloged by metagenomic analysis (Pearson correlation coefficient r=0.0708, p=0.83; Supplemental Table S2), but did observe a significant positive correlation between the proportion of reads mapping to the TB genome and the proportion that were unambiguously classified as TB by our high specificity approach (Pearson correlation coefficient r= 0.6727, p=0.017; Supplemental Table S2). Taken together, these findings indicate that the contributions of non-TB organisms to the analysis are minor, and that the reads being mapped to the TB genome are largely attributable to TB-derived cfDNA.

While the length distribution of human reads by this approach was consistent with our earlier results (Figure 1C), with the most abundant human cfDNA fragment length ranging from 29–53 bp across samples, cfDNA fragments mapping to the TB genome were substantially shorter than previously indicated (Figure 1D). The abundance of TB urine cfDNA increased exponentially with decreasing fragment size (Figure 1D) and showed a peak fragment length of ≤19 bp, the minimal size that is detectable by our analysis, in most samples (8/10) (Figure 1E). The median fragment lengths for TB-derived urine cfDNA (≤19–52 bp) remained significantly shorter than for human urine cfDNA (42–92 bp) (p=0.002 by Wilcoxon matched pairs test, Figure 1E).

These mapped reads were used for subsequent analyses.

### Distribution of TB-derived reads across the genome

In TB-positive participants, cfDNA reads mapping to the TB reference genome showed low, but relatively uniform coverage across the length of the genome (Figure 2A). Notably, for most samples the rRNA gene locus (positions 1471846 to 1477013 bp), evidenced increased read coverage relative to the rest of the TB genome, despite there being a single copy of this locus carried by TB (26). As rRNA encodes an essential gene that is highly conserved across bacterial taxa (27), these data suggest that short reads derived from other organisms present in patient specimens may infrequently map to the TB genome at specific sequence contexts.

**Figure 2:**
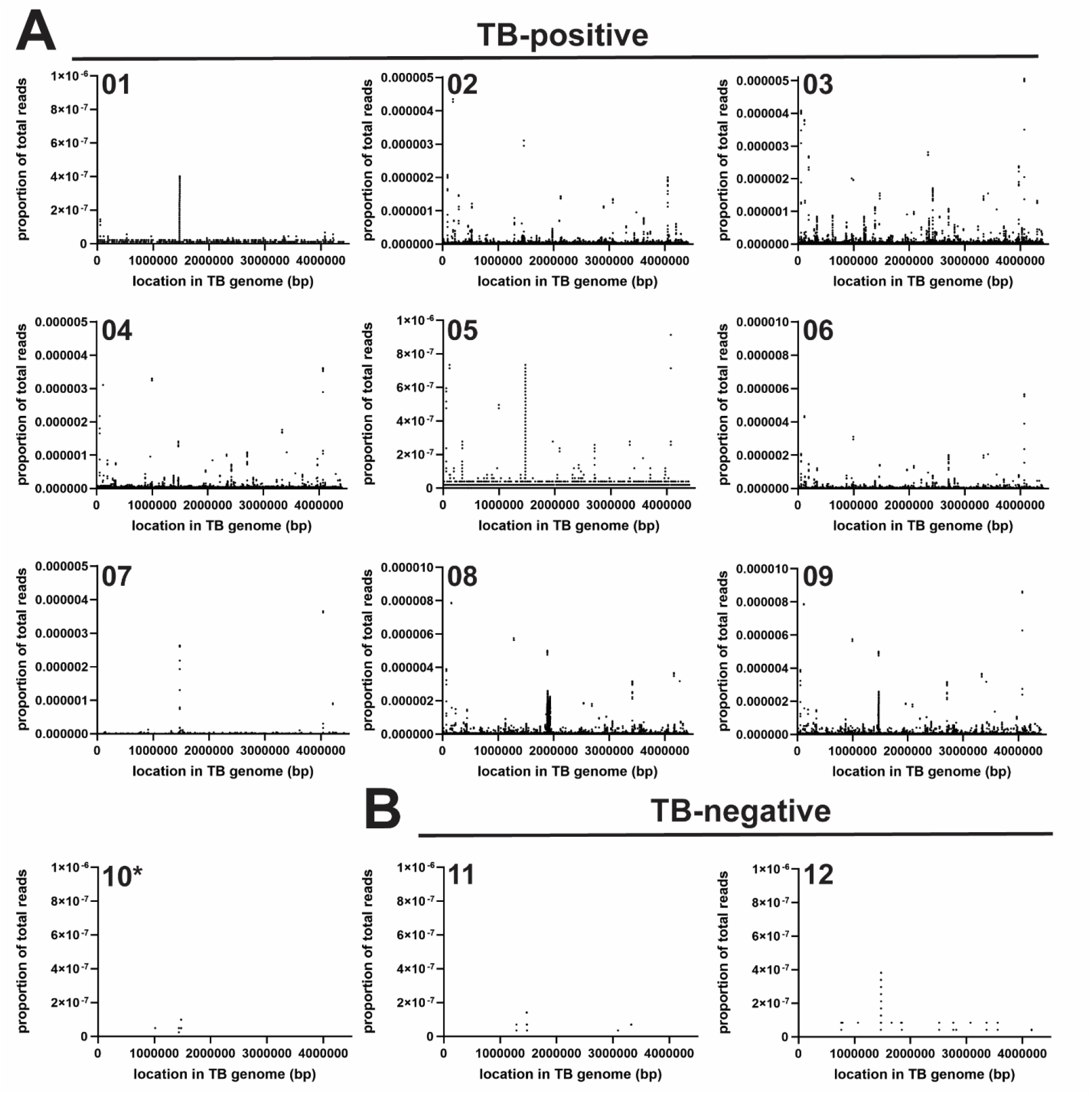
Coverage of the TB genome in urine cfDNA. **(A)** Density of reads mapped to the TB genome in ten samples from TB-positive participants. *Sample 10 had no TB cfDNA detectable by IS6110 qPCR, but TB-specific cfDNA was detectable by sequencing and confirmed by metagenomic classification analysis (kraken2). **(B)** Density of reads mapped to the TB genome in two samples from TB-negative participants.

### Multicopy elements in the TB genome as diagnostic targets for urine cfDNA

Species-specific multicopy elements are attractive targets for diagnostic testing because they provide both specificity and an inherent level of signal amplification. To evaluate the potential of two known multicopy genomic elements as potential urine cfDNA diagnostic targets, we analyzed the relative abundance of reads mapping to two insertion sequences (IS6110 and IS1081) present in the TB genome (28, 29).

cfDNA fragments derived from IS6110 and IS1081 cfDNA were detected by NGS in 6/9 and 5/9 specimens with TB cfDNA detectable by IS6110 qPCR, respectively (Table 2). In cases where these sequences were identified, they were found with greater abundance than reads from other regions of the TB genome. The average fold overrepresentation of IS6110 relative to the average sequencing depth for the remainder of the TB genome was 8.1 (range 1.3–17.1), whereas that observed for IS1081 was 4.6 (range 1.7–8.5). These values correspond roughly to the expected count of each element per genome. IS6110 is present at variable copy number (0–25 copies) across MTB complex strains (30), while IS1081 is present at a more stable copy number (5–6 copies) (31). We were not able to examine reads mapping to a third multicopy element used in TB studies, the direct repeat (DR) region (32), given the repetitive nature of the element and the short length of its constituent repeat sequence (36 bp, present at an estimated 14–63 copies per genome), which prevented reliable read mapping.

**Table 2:** Relative abundance of multicopy elements IS6110 and IS1081 in urine cfDNA.

| Sample ID | TB status | cfDNA status (IS6110 qPCR) | IS6110 fold overrepresentation<sup>a</sup> | IS1081 fold overrepresentation<sup>a</sup> |
| --- | --- | --- | --- | --- |
| 01 | Positive | Positive | 0 | 0 |
| 02 | Positive | Positive | 0 | 0 |
| 03 | Positive | Positive | 17.0 | 2.52 |
| 04 | Positive | Positive | 1.27 | 0 |
| 05 | Positive | Positive | 3.30 | 6.78 |
| 06 | Positive | Positive | 3.68 | 3.62 |
| 07 | Positive | Positive | 17.1 | 8.48 |
| 08 | Positive | Positive | 6.34 | 1.72 |
| 09 | Positive | Positive | 0 | 0 |
| 10 | Positive | Negative | 0 | 0 |
| 11 | Negative | Negative | 0 | 0 |
| 12 | Negative | Negative | 0 | 0 |

<sup>a</sup> Measured as average sequencing depth at the target region normalized to the average sequencing depth across the remainder of the TB genome.

## DISCUSSION

In this study, we present an in-depth, unbiased characterization of TB urine cfDNA using NGS. To most comprehensively characterize the range of fragments occurring in this analyte, we selected DNA extraction and sequencing library preparation methods specifically for their demonstrated effectiveness for short DNA fragments. We have previously shown in a comparison of urine cfDNA purification methods that Q Sepharose extraction, which pre-concentrates urine cfDNA using anion exchange resin prior to desalting on a silica spin column (15), has a high rate of recovery (>70%) for spiked DNA down to at least 40 bp in length (16). Recovery by that method is reduced to <10%, but is still measurable, for DNA as short as 25 bp in length (16). Similarly, single-stranded NGS library preparation (11, 12) has been shown to improve recovery of <100 bp cfDNA, having a lower reported limit of 40–60 bp (13). In this application, we have further extended the lower range of detection for this approach by retaining all library fragments generated, which increased our sensitivity for low molecular weight DNA fragments at the expense of sequencing an increased proportion of synthetic, noncontributory fragments resulting from self-ligated sequencing adaptor molecules (measured at 9% to 20% of total reads generated per specimen).

Previous NGS studies that characterized the fragment length distribution of human genomic cfDNA have reported peak fragment lengths of approximately 50–100 bp (14, 22, 23). In contrast, our study demonstrably improved recovery of short cfDNA fragments and revealed a previously undetectable fraction of human genomic cfDNA in urine having a peak fragment length of 29–53 bp, which was observed in all samples. The differences between our methodology and protocols employed previously were most noticeable for the shortest fragments, with representation of <50 bp fragments in our study dramatically increased relative to earlier work that did not use single-stranded library preparation methods (22–24) or which used single-stranded library preparation in conjunction with a DNA extraction method less able to efficiently recover short DNA fragments (Qiagen Circulating Nucleic Acid Kit) (14).

Our results provide evidence that TB cfDNA in urine is extensively fragmented, significantly more so than human genomic cfDNA (Figure 1). Consequently, minimizing the length of sequences targeted for TB urine cfDNA diagnostic assays will be critical to maximizing their sensitivity. Decreasing the minimum detectable target length improves detection sensitivity for fragmented cfDNA (15, 33, 34) and has been a priority during the recent development of TB urine cfDNA assays (5–8). Previously reported TB urine cfDNA assays targeted, at the shortest, amplicons of 38–40 bp (6–8). Decreasing PCR amplicon length from 49 bp to 39 bp resulted in greater than 10-fold increase in detected TB cfDNA (34). Until now, the extent to which further decreases in target length may improve sensitivity has been unclear. Our results suggest that even small, incremental decreases in target length may have a disproportionate impact on detection of TB urine cfDNA, which is highly fragmented to a peak size of ≤19 bp and increases in abundance exponentially as fragment size decreases. To improve the clinical sensitivity of TB urine cfDNA assays, both sample preparation and amplification methods having maximal efficiency for very short fragments are needed. For example, recent work in our lab demonstrated that sequence-specific purification improves recovery of short cfDNA relative to conventional silica-based extraction and increases the clinical sensitivity of TB diagnosis from urine cfDNA (8, 35). Ultrashort PCR using a stem-loop primer may be an attractive strategy for amplification of fragments too short for conventional PCR (15). Our results, in concert with those of a previous study (10), suggest that targeting multicopy genomic elements (*e*.*g*., IS6110, IS1081) is likely a more promising strategy than identification of *de novo* cfDNA targets.

Our study has several limitations. First, we sequenced cfDNA only from patients living with HIV. Detection sensitivity for TB urine cfDNA is similar in HIV-positive and HIV-negative participants (7, 8), but it remains unclear if there may be differences in cfDNA fragmentation patterns across these two populations. Second, owing to the requirements for high sequencing depths and attendant sequencing costs, the number of specimens analyzed in this study are necessarily limited. Third, despite the improvements in short cfDNA fragment recovery using a combination of Q Sepharose DNA extraction and single-stranded library preparation, our methods are unable to reliably interrogate the shortest cfDNA fragments. We expect that efficiency of fragment recovery begins to decrease below 40 bp (16), and due to the nature of sequence read mapping algorithms, it is not possible to reliably map the origin or sequence reads below a specified seed length (here, 19 bp). Moreover, the shorter the fragment length, the less probable it is that the read will map confidently to its target (36). Considering these limitations, the true frequency of cfDNA less than 40 bp in length, whether originating from human or TB, is likely even greater than registered by our analysis. The fragment length distribution of cfDNA should be interpreted with this in mind. Fourth, many of the cfDNA molecules recoverable by our methods are so short that they cannot be uniquely classified as belonging to TB. As a consequence, we cannot demonstrate directly that all of the smallest read fragments we mapped to TB derive from derive from that organism, although accessory evidence is consistent with that conclusion.

In summary, accurate characterization of urine cfDNA using NGS provides critical insight into its validation as a biomarker for TB. Our findings, in particular the discovery that TB cfDNA is substantially shorter than human genomic cfDNA, will help inform the development of improved assays for TB diagnosis from urine cfDNA. The large potential sensitivity benefit to be gained by targeting <40 bp TB cfDNA motivates continued prioritization of both sample preparation and amplification methods designed for short fragments, although this will need to be balanced against reduced specificity accompanied by interrogating shorter nucleotide fragments. A sensitive molecular assay targeting urine cfDNA, rather than sputum, would considerably contribute to improving sample accessibility and diagnostic yield and has the potential to advance availability of rapid TB diagnostics across underserved patient populations. In addition, the combination of Q Sepharose DNA extraction and single-stranded library preparation will likely prove useful for other applications and contexts where the analysis of highly fragmented forms of DNA is necessary.

## Data Availability

All relevant data are in the manuscript, its supporting information files, and in the NCBI Sequence Read Archive (SRA) under accession PRJNA725220.

## ACKNOWLEDGEMENTS

We are grateful to study participants for their contributions to this research and thank the KwaZulu-Natal Department of Health and staff of Edendale Hospital for their partnership. Research reported in this publication was funded by the Brotman Baty Institute for Precision Medicine Catalytic Collaborations Program and the Bill and Melinda Gates Foundation under award number OPP1213054. A.O. was supported by funding from the National Science Foundation Graduate Research Fellowship Program. The funders had no role in study design, data collection and interpretation, or the decision to submit the work for publication.

