## Supplemental Figure S1 for "Unbiased sequencing of *Mycobacterium tuberculosis* urinary cell-free DNA reveals extremely short fragment lengths"

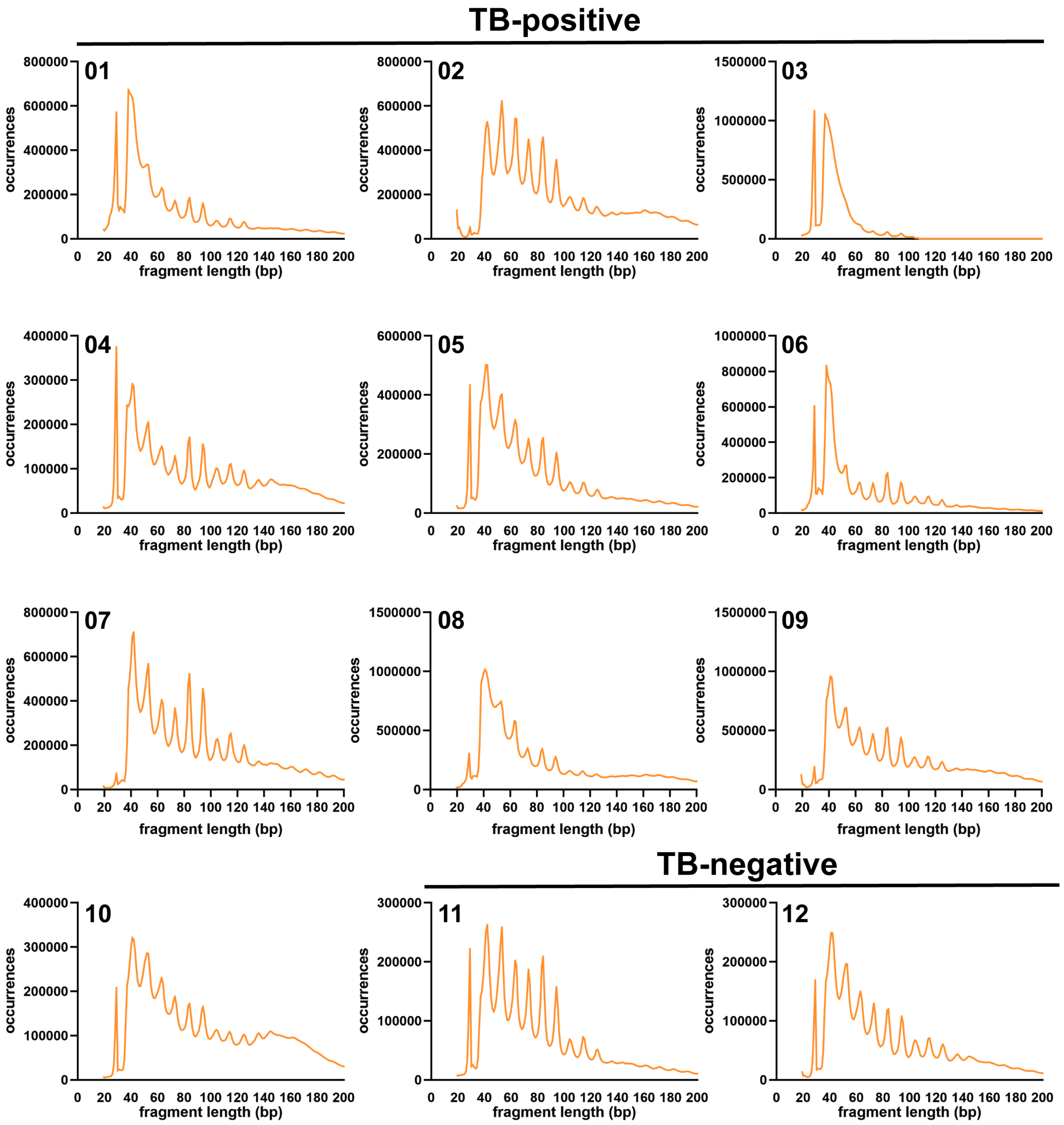


**Supplemental Fig. S1: Fragment length distribution of cfDNA mapped to the human genome in each sample.**
