## Supplemental Figure S2 for "Unbiased sequencing of *Mycobacterium tuberculosis* urinary cell-free DNA reveals extremely short fragment lengths"

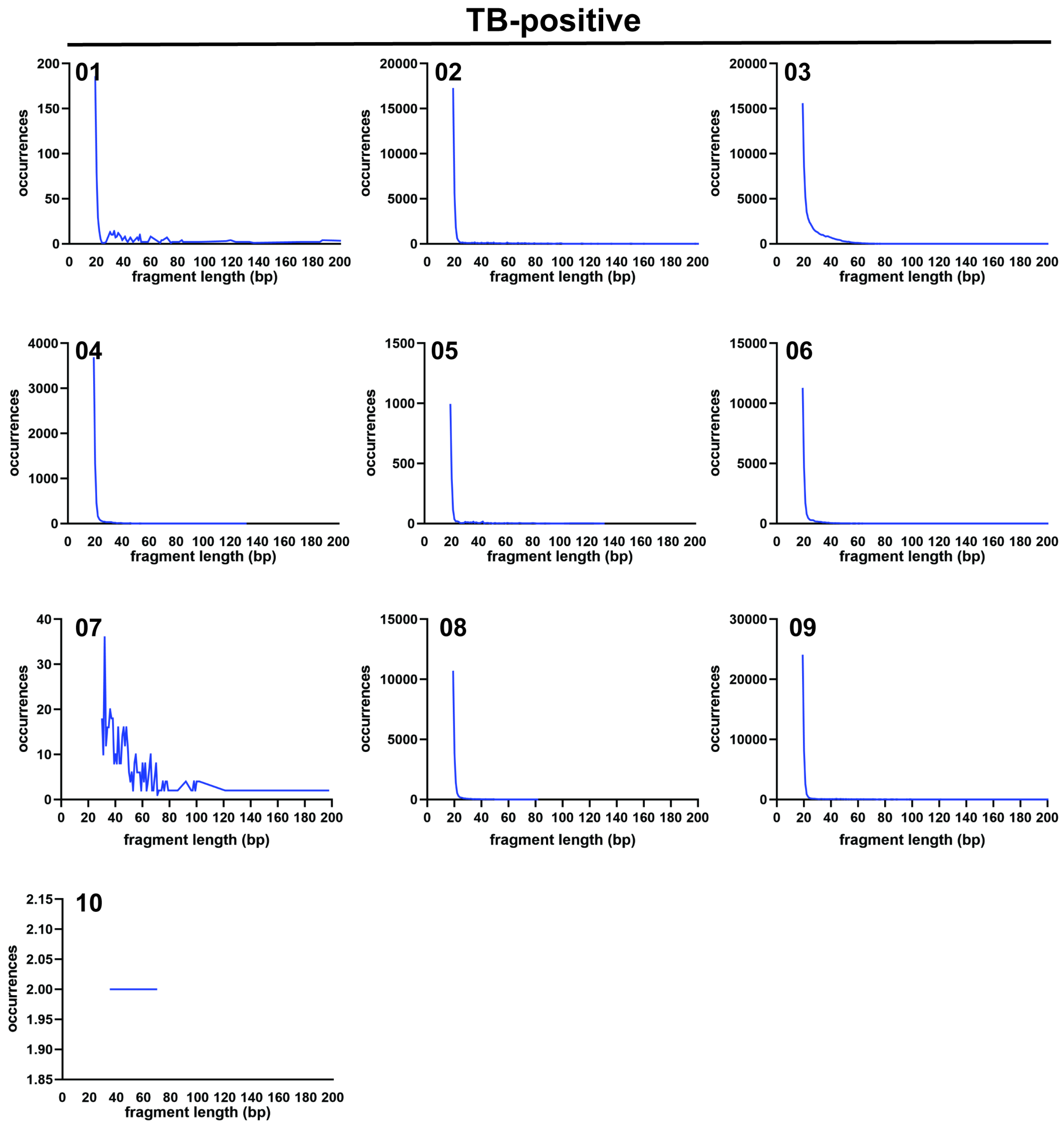


**Supplemental Fig. S2: Fragment length distribution of cfDNA mapped to the TB genome in each sample.** Sample 10 had a low number of reads that mapped to the TB genome, but was confirmed to contain reads specific to TB by metagenomic classification. Neither TB-negative sample (Sample 11 or Sample 12, excluded here) contained reads specific to TB by metagenomic classification.
